# Knowledge and attitudes towards thalassemia among university students in Bangladesh

**DOI:** 10.1101/2022.09.19.22280125

**Authors:** Jubayer Hossain, Syeda Tasneem Towhid, Sabia Sultana, Sumaiya Akter Mukta, Rubaiya Gulshan, Sharif Miah

**Author notes:** Corresponding author Dr. Syeda Tasneem Towhid.

## Abstract

**Background:** Thalassemia is the most common congenital single-gene condition. It is marked by a lack of or reduced synthesis of either the alpha- or beta-globin chains and passed down from parents to offspring. This study aimed to determine how healthy students in Bangladeshi public universities were aware of thalassemia and how they felt about it.

**Methods:** A cross-sectional descriptive online survey was conducted on public university students in Bangladesh using a structured questionnaire between June and November 2020. Students completed structured questionnaires consisting of three sections: demographic information, ten multiple-choice knowledge questions rated on a scale of 0 to 10, and ten attitude questions. The data were analyzed using Python. Descriptive statistics methods such as frequencies and percentages were used to present data.

**Results:** A total of 681 students participated in the online survey. The average age of the respondents in this study was 21.97 years, with a standard deviation of 2.9. Most of the students, 611(89.72%), had heard about thalassemia. Only 248 (36.42%) of the students had a fair understanding of thalassemia, with 138 (22.62%) being male and 110 (18.03%) being female (P-value = 0.0819). Students’ knowledge level did not differ significantly by gender (P-value = 0.0819), marital status (P-value = 0.2281), or academic year (P-value = 0.4619), but there was a substantial variation by field of study (0.0042). However, 478 (78.36%) participants showed a positive attitude toward “Premarital Screening” to prevent thalassemia.

**Conclusions:** In Bangladesh, where the rate of family marriage is high, it is recommended that university students and the general public engage in long-term, goal-oriented prevention measures. These initiatives would provide crucial information and increase people’s awareness of thalassemia when married and after becoming parents, reducing the impacts of illnesses.

## Introduction

Thalassemia is the most prevalent congenital single-gene disorder, characterized by a lack of or decreased synthesis of either the alpha- or beta-globin chains and transmitted from parents to children. Thalassemia is a genetic blood disorder in which most patients cannot produce red blood cells and must depend on blood transfusions for the rest of their lives (1). Food, sleep, living, and breathing do not affect it. Although thalassemia is a preventable disease, it’s a high prevalence, and the lack of cure has made it a global health concern (2). The most prevalent clinical signs of thalassemia, classified as alpha, beta, gamma, or delta, are alpha and beta-thalassemia. Beta-thalassemia is organized into three clinical severity and inheritance pattern categories: severe, minor, and moderate.

Thalassemia is highly preventable in Southeast Asia, the Indian subcontinent, the Mediterranean, Middle Asia, Central Asia, and West Africa(2). As a result of migration to non-endemic areas, thalassemia spread across Europe and North America. It is a disease that can be avoided, as countries like Italy, Greece, and Cyprus have proved (3). Repeated blood transfusions, and symptomatic or permanent therapies, such as bone marrow transplantation, are out of reach for most patients in developing nations(4). Although no cure exists, prevention is possible, implementable, and successful in some areas(5). Every year, over 14,000 thalassemia children are born in our country, with the condition affecting 10% of the general population and more than 30% of the tribal population (6).

The number of thalassemia patients increases due to a lack of understanding, poor educational activities, lack of awareness due to low literacy rates, socioeconomic causes, religious choices, and cultural limitations(3). Thalassemia can be reduced through preventive health services, awareness, screening, prenuptial genetic counseling, and prenatal diagnosis in consanguineous marriages. Complete blood counts (CBC) are combined with a single tube osmotic fragility test (SOFT), and hemoglobin electrophoresis is used to corroborate the results. In thalassemia testing, a chorionic villus sample is also used (7).

Because today’s youth will be the parents of tomorrow, their knowledge and attitude regarding thalassemia can aid in the fight against the disease. As a result, with sufficient knowledge and comprehension of conditions, they should be able to overcome the adverse effects on their marriage(8). This study aims to determine the degree of awareness and attitudes toward thalassemia among public university students in Bangladesh.

Thus, the long-term goal of this study is to understand how awareness campaigns could be more effective in premarital screening and genetic counseling in preventing thalassemia.

## Methods

### Study design

A cross-sectional descriptive online survey was designed, and the response was collected between June to November 2020 among public university students in Bangladesh. Graduate and undergraduate students from several public universities and students from various fields of study such as arts and humanities, science, business, and social science were included in the survey. Participants who used social media were also included (Facebook, Instagram, WhatsApp, etc.). Incomplete surveys and those who refused to participate in the study were disqualified. There were no incentives or rewards offered to participants, and all responses were anonymous. The study participants were chosen using the snowball sampling technique.

### Instruments

The research team devised a self-reported questionnaire after conducting a thorough literature review (1,2,4,7,9–14) on individuals’ thalassemia knowledge, attitudes, and practices. The questionnaire was in English. A brief explanation of the research background, aims, eligibility requirements, confidentiality statement, who can participate, and the online consent form appeared on the first page of the questionnaire. Students then completed structured surveys with demographic information, an awareness section with 10 multiple-choice questions, and an attitude section with 10 questions.

### Data collection

Using an online survey tool (Google Forms), the researchers gathered data from social media networks (Facebook, WhatsApp). Participants were asked if they were willing to participate in the study via “yes/no” responses. They were provided access to the entire questionnaire if they replied “yes.” Otherwise, a blank survey form was submitted right away. 685 people completed the online survey, with 660 included in the final analysis after quality control and manual checks to eliminate incorrect surveys.

### Data management and analysis

Data was downloaded into Microsoft Excel from Google Sheets and was processed for labeling categories and missing data. The study involved 681 students; however, only 660 responses were included in the final analysis. The cleansed data were analyzed using descriptive statistics such as counts, percentages, and mean age computation. The Chi-Square test was used to investigate the relationship between gender, the field of study, education level, and thalassemia knowledge and attitudes. A statistically significant P-value of 0.05 was used. R (version 4.0.2) and Rstudio (version 1.3.1056) for Windows were used for all analyses.

### Scoring of knowledge level

This study’s participants were asked ten knowledge-related questions, which were graded on a scale of 0 to 10. Google forms auto-scoring methods were used to determine the knowledge scores. Poor, average, and good knowledge are the three levels of knowledge (less than 33%, between 33% to 67%, and more than 67% of questions were correctly answered, respectively.).

## Results

An online survey on thalassemia awareness depicted the state of knowledge and attitude towards the dreaded disease and its management among university students in Bangladesh.

Table 1 shows the distribution of responses (n=660) to queries about thalassemia source information. 611 students (89.72%) had heard of thalassemia before. On the other hand, 70 students (10.28%) were clueless. Electronic media (45.41%), friends and family (41.80 %), healthcare professionals (18.20 %), print media (25.90%), seminars and lectures (25.74%), and other sources were the primary sources of information for these 611 students (72%).

**Table 1.** Distribution of (n = 660) responses to questions about the information source for thalassemia.

| Background Information | N (%) |
| --- | --- |
| Have you previously heard about thalassemia? |  |
| Yes | 611 (89.72) |
| No | 70 (10.28) |
| If yes, what information sources did you hear about thalassemia? |  |
| Electronic Media | 277 (45.41) |
| Friends and Family | 255 (41.80) |
| Healthcare Professionals | 111 (18.20) |
| Print Media | 158 (25.90) |
| Seminar & Lectures | 157 (25.74) |
| Others | 41 (6.72) |

Table 2 summarizes the results of sociodemographic variables. The participants included 336 males (55.08%) and 274 females (44.92%). There were 562 unmarried students (92.13%) and 48 married students (7.87%). In this study, 414 students (67.87%) majored in science, 72 students (11.80%) majored in business, 69 students (11.31%) majored in humanities, and 55 students (9.02%) majored in social science. Around 240 students (39.34%) were in their first year, 114 (18.69%) in their second year, and 107 (17.54%), 101 (16.56%), and 48 (7.87%) in their third, fourth, and master’s year, respectively.

**Table 2.** Demographic characteristics of participants and the proportion of the participants who heard about thalassemia.

| Variables | Response who heard about thalassemia n (%) |
| --- | --- |
| Sex |  |
| Male | 336 (55.08) |
| Female | 274 (44.92) |
| Marital Status |  |
| Unmarried | 562 (92.13) |
| Married | 48 (7.87) |
| Field of Study |  |
| Science | 414 (67.87) |
| Business | 72 (11.80) |
| Arts and Humanities | 69 (11.31) |
| Social science | 55 (9.02) |
| Year of Study |  |
| 1st year | 240 (39.34) |
| 2nd year | 114 (18.69) |
| 4th year | 107 (17.54) |
| 3rd year | 101 (16.56) |
| Masters | 48 (7.87) |

Table 3 shows the associations between sociodemographic factors and thalassemia knowledge among students. The academics had quite different perspectives on thalassemia. Science students had the highest knowledge level, with 21 (1.97%), followed by social and business students, with 22 (3.61%) and 19 (3.11%), respectively. Many science students (7.54% and 29.84%) had weak and average knowledge levels. Students in the arts and humanities, social science, and business faculties all had low and intermediate knowledge levels that were nearly identical (1.97%, 2.13%, 2.46%). P values for sociodemographic factors differ between students. For variables such as sex, marital status, the field of study, and year of study, P values of 0.0819 %, 0.2281 %, 0.0042 %, and 0.4619 % were discovered.

**Table 3.** Demographics of the participants in the survey for Thalassemia Awareness in Bangladesh.

| Variables | Level of Knowledge |  |  |  |
| --- | --- | --- | --- | --- |
|  | Weak | Average | Good | P-value |
| Sex |  |  |  |  |
| Male | 56 (9.18) | 142 (23.28) | 138 (22.62) | 0.0819 |
| Female | 30 (4.92) | 134 (21.97) | 110 (18.03) |  |
| Marital Status |  |  |  |  |
| Married | 3 (0.49) | 22 (3.61) | 23 (3.77) | 0.2281 |
| Unmarried | 83 (13.61) | 254 (41.64) | 225 (36.89) |  |
| Field of Study |  |  |  |  |
| Science | 46 (7.54) | 182 (29.84) | 186 (30.49) | 0.0042 |
| Arts &<br>Humanities | 12 (1.97) | 36 (5.90) | 21 (3.44) |  |
| Social Science | 13 (2.13) | 20 (3.28) | 22 (3.61) |  |
| Business | 15 (2.46) | 38 (6.23) | 19 (3.11) |  |
| Year of Study |  |  |  |  |
| 1st-year | 31 (5.08) | 112 (18.36) | 97 (15.90) | 0.4619 |
| 2nd-year | 19 (3.11) | 56 (9.18) | 39 (6.39) |  |
| 3rd-year | 17 (2.79) | 47 (7.70) | 37 (6.07) |  |
| 4th-year | 13 (2.13) | 44 (7.21) | 50 (8.20) |  |
| Masters | 6 (0.98) | 17 (2.79) | 25 (4.10) |  |

Table 4 shows the opinions of students about thalassemia (n=611). The percentage of pupils increased since they didn’t know anything about thalassemia. The kids erroneously answered the majority of the thalassemia questions. Only 35.25% of students would accept a thalassemic association, while 29.67 % were against and 35.08 % were neutral. Nearly 77.05% stated they would consult a wedding planner before getting married. Only 78.36% favored blood testing before marriage to avoid the birth of a thalassemic kid, while 4.92% were opposed, and 16.72 % were undecided. Only 90.98% said thalassemia education should be available to the general public. Most people (91.48%) said they wanted to donate their bone marrow to their relatives. About 80.82 % felt that taking part in a “Thalassemia Prevention Program” if one is offered is a better approach to avoiding thalassemia, while 3.11% were unsure.

**Table 4.**
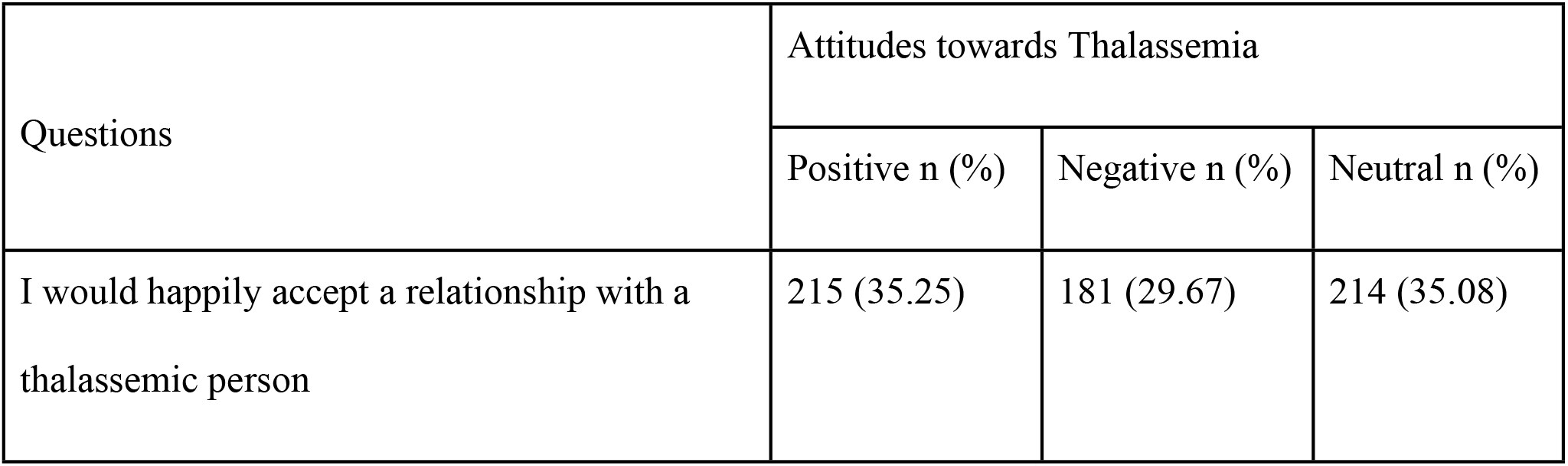
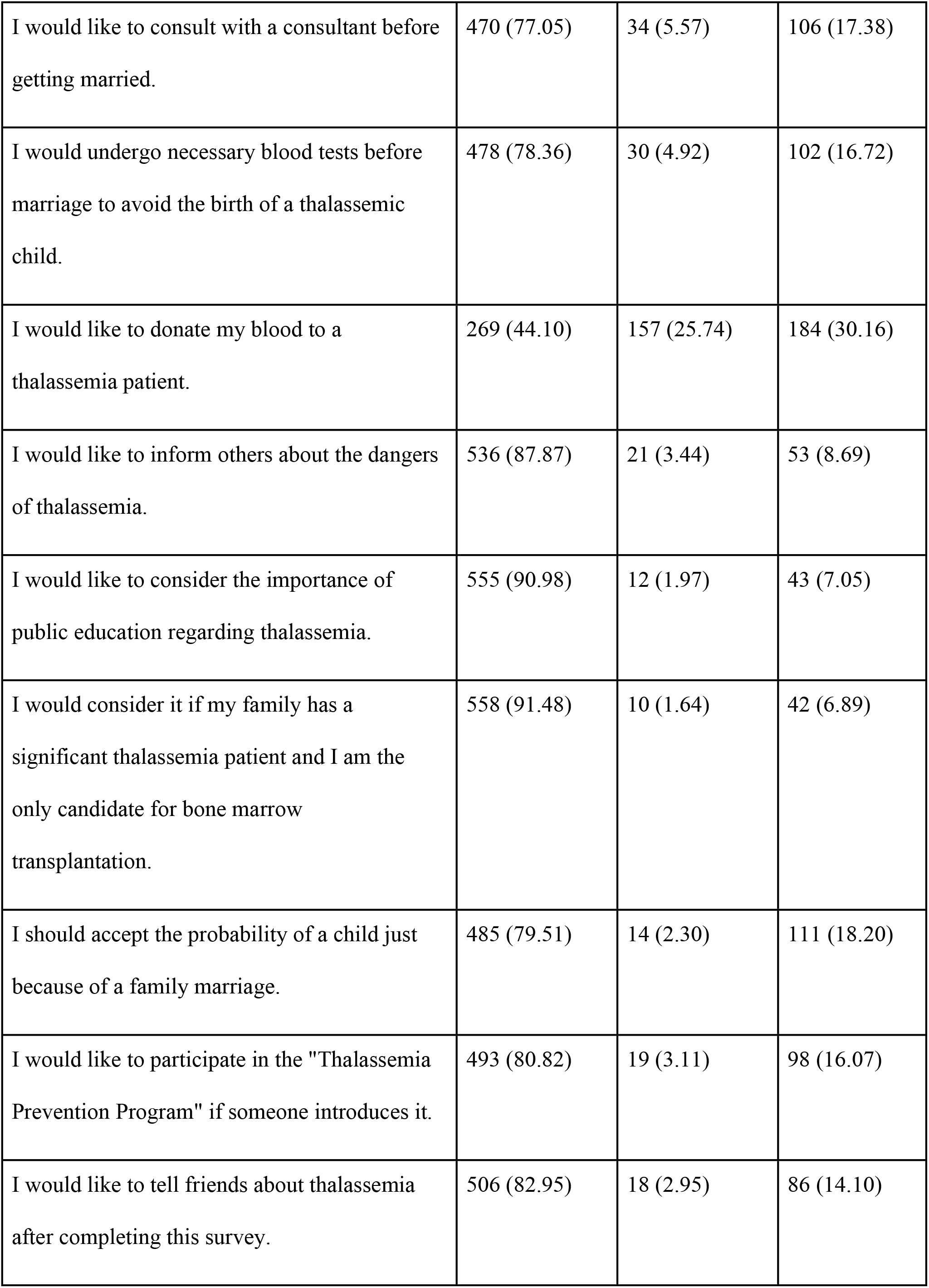
Attitudes towards thalassemia awareness among participants (n = 611)

## Discussion

This study aimed to see how well Bangladeshi public university students understood thalassemia and how they felt about it. Our research helped to fill knowledge gaps about thalassemia. Thalassemia is a genetic hemoglobin synthesis disorder that causes anemia and insufficient erythropoiesis due to faulty and unequal globin chain production.

The most effective treatment for this sickness is bone marrow transplantation; however, most people cannot afford it owing to financial constraints. Significant resources have been focused on illness prevention in wealthy countries, such as finding thalassemia carriers and marriage counseling. Such preventive methods facilitate diagnosis and treatment and the management of the current sickness burden. Although there is no cure, prevention is possible and has been done successfully in various countries. 89.72 percent of the students interviewed had heard of thalassemia, according to our data.

Our major findings reveal that thalassemia awareness is low, with one-third of the participants (10.28%) having never heard of the disease. Even many who had heard of thalassemia lacked a basic understanding of the disease. The most common sources of information for people who had heard of thalassemia were electronic media (41.41 %), friends and relatives (41.80%), print media (25.90%), seminars, and lectures (25.74 percent%), and others (6.72%). Physicians performed a small impact on their patients’ comprehension of thalassemia (18.20%). This could be because the publications and awareness drives are centered around International Thalassemia Day instead of throughout the year. The most startling finding of our research was that of the four faculties assessed. Science students had the highest level of knowledge (67.87%), followed by business (11.80%), arts and humanities (11.31%), and social science students (11.31%) (Table 2). (9.02%). In this study, 35.25%of students had positive attitudes toward thalassemia and wanted to embrace a connection with a thalassemic patient joyfully, while 29.67% had negative attitudes, and 35.08% had neutral attitudes (Table 4). A negative attitude could stem from ignorance and a lack of empathy.

Regarding the relevance of a blood test before marriage to prevent the birth of a thalassemic child, 78.36% of students agreed, 4.92% disagreed, and 16.72% were undecided (Table 4). Nearly 77.05% of students decided to visit a doctor before marrying to avoid thalassemia. In our poll, 44.10 % of respondents stated they would donate blood to a patient with Thalassemia (Table 4). Around 79.51% believed they should expect to have a child simply because they married into a family with a history of thalassemia, whereas 2.30% strongly disagreed and 18.20% were undecided (Table 4). Approximately 87.87 percent of students stated they would inform others about thalassemia’s dangers. In the context of thalassemia, about 90.98% were aware of the importance of public education (Table 4). This is supported by the fact that 80.82% of students stated they would participate in a “Thalassemia prevention program” if one was offered, whereas 16.07% said they were neutral. After completing the study, however, 82.95% of individuals who took part agreed to inform their friends about thalassemia.

Like lower-middle-income countries, Bangladesh doesn’t have medical insurance or other social security/safety net regarding high medical expenses (10). Access to cutting-edge interventions such as gene therapy or bone marrow transplant is minimal (10). The efforts to minimize preventable conditions such as thalassemia are centered around awareness drives once a year, which is too little for a population with a high density of heritable hemoglobinopathy(9). Up to 10% of the Bangladeshi population could be Thalassemia carriers, with 15000 new Thalassemia-positive births annually. The cost of treatment comes to BDT 20,000 (USD 230) monthly, whereas the GDP per capita is USD 1969 (Regional Desk Review of South East Asia, WHO 2021). Treatment provisions are also limited, with one specialized hospital and several district hospitals offering transfusion and iron chelation/hydroxyurea medication (15). The better option for fighting thalassemia is the reduction of Thalassemia births with premarital, prenatal, and target screening (10). Cypress has reduced Thalassemia births to a negligible number over the last decade through an awareness campaign and premarital screening (13). This study was an incremental step toward assaying the knowledge gap to devise an action plan that effectively disseminates information within the shortest time.

Our research has some limitations: for starters, all data for this study was not collected through face-to-face interviews because of COVID-19 pandemic situations, and their responses may be inaccurate because of social desirability factors. Second, we did not measure the level of awareness of each student individually. Additional research is needed to discover differences in knowledge based on gender, education, marital status, and awareness level. Due to a lack of time, this study focused only on the non-medical community’s management. Finally, the self-reported questionnaire may have resulted in response bias because the questionnaire was not checked and was based on past research. Thalassemia is becoming a rising public health concern in Bangladesh. The study highlights the need to educate individuals about the dangers of consanguineous marriage, which is the most significant part of preventing thalassemia.

Additionally, thalassemia testing for all couples should be made accessible and necessary. A thalassemia prevention program should be designed and interventions deployed throughout Bangladesh to eradicate the disease. According to this curriculum, students remembered their carrier status and took prophylactic measures.

## Conclusion

Finally, this study accomplishes the investigation’s purpose of analyzing thalassemia knowledge and attitudes among Bangladeshi public university students. They had inadequate knowledge and attitudes about thalassemia, according to our findings. Most pupils were vaguely familiar with the disease or knew little about it. The majority of these students had no idea they had a hereditary condition. Furthermore, kids’ levels of knowledge and awareness can differ substantially depending on their socioeconomic situation. Large-scale experiments with a diverse study population are required to confirm the findings. Because they impact students’ minds, the mass media (print, television, and radio) must contribute to thalassemia awareness.

Despite a countrywide mandated thalassemia premarital examination screening program, the incidence of high-risk thalassemia has not decreased much. On the other hand, a far larger scale of health education about thalassemia awareness and prevention is required. The study recommends hosting seminars and public lectures for thalassemic families and the general public to understand thalassemia disease and prevention better. People will recognize the disease’s importance in their daily lives. Therefore, raising awareness through community participation will be beneficial. As a result, implementing a complete prevention program comprising premarital counseling, genetic testing, prenatal screening, and community-based awareness programs may help reduce the prevalence and incidence. This paper will substantially contribute to thalassemia prevention in resource-constrained nations like Bangladesh.

## Data Availability

The dataset used and analyzed during the study is available from the corresponding author and ethical review committee.

## Ethics approval and consent to participate

This study adhered to the most significant ethical standards imaginable, and participants gave informed consent. Informed consent was also obtained from the guardian of each participant under 18 years of age. The Helsinki Declaration was observed in all procedures. Anonymity and confidentiality were maintained. Anonymity and confidentiality were maintained. We obtained ethical approval for this study from the Ethical Review Committee of CHIRAL Bangladesh (Reference No: CHIBAN24MAY2020-0001)

## Availability of data and materials

The datasets used and analysed during the current study are available from the corresponding author on reasonable request.

## Competing interests

The authors declare that they have no potential conflict of interest in publishing this research output.

## Funding

The authors did not receive funding from any individuals or organizations for this study.

## Authors’ contributions

Conceptualization: Md. Jubayer Hossain

Data curation: Md. Jubayer Hossain, Sumaiya Akter Mukta, Rubaiya Gulshan, Md. Sharif Miah

Formal analysis: Md. Jubayer Hossain, Dr. Syeda Tasneem Towhid

Investigation: Md. Jubayer Hossain, Dr. Syeda Tasneem Towhid

Methodology: Md. Jubayer Hossain

Project Administration: Md. Jubayer Hossain, Dr. Syeda Tasneem Towhid, Sabia Sultana

Resources: Md. Jubayer Hossain

Software: Md. Jubayer Hossain

Supervision: Dr. Syeda Tasneem Towhid, Sabia Sultana

Validation: Md. Jubayer Hossain, Dr. Syeda Tasneem Towhid

Writing – Original Draft Preparation: Md. Jubayer Hossain, Dr. Syeda Tasneem Towhid, Sabia Sultana, Sumaiya Akter Mukta, Rubaiya Gulshan, Md. Sharif Miah

Writing – Review & Editing: Md. Jubayer Hossain, Dr. Syeda Tasneem Towhid, Sabia Sultana, Sumaiya Akter Mukta, Rubaiya Gulshan, Md. Sharif Miah

All authors have read and approved the manuscript.

## Acknowledgments

Firstly, the authors would like to express the most profound gratitude to all respondents who participated in this study. The authors would like to thank all research assistants:Rasel Hossain, Shumon Pramanik, Farhan Shahriar Pranto, Rifat Ara Yasmin (Rajmoni), Rubaiya Gulshan Meem, Sharif Mia, Sumaiya Akter Mukta, Wahid Arafat, Rumaiya Sazneen Moni, Rahul Roy Aviraj, Sathi Paul, Nargees Akter, Spencer Mark, Depro Das, Durdana Priom, Fatema Mehejabin Trina, Sajib Kumar, Momin Ali, Md. Kamrul Islam, Muhibullah Shahjahan, Nayem Reaid, Nowshin Sharmile, Farida Akter Farzana, Esmot Ara Shoshi, Md Mehedi Hasan, Iffat Noor, Md Reon Tanvir Anando, Abir Hossain, Akash Kumar, Rafia Nusrat Prome, for their supports during the study periods.

## Notes

### Competing Interest Statement

The authors have declared no competing interest.

### Funding Statement

The author(s) received no specific funding for this work.

### Author Declarations

This study adhered to the most significant ethical standards imaginable, and participants gave informed consent. The Helsinki Declaration was observed in all procedures. Anonymity and confidentiality were maintained. Anonymity and confidentiality were maintained. We obtained ethical approval for this study from the Ethical Review Committee of CHIRAL Bangladesh (Reference No: CHIBAN24MAY2020-0001)

## References

1. Ishfaq K. The Knowledge of Parents Having Thalassemia Child. Isra Med J. 2017;8(June 2016):79–83.

2. Alam N-E-, Islam S, Suries U, Binti R, Islam M, Akter S, et al. Public Perceptions and Attitudes of Bangladeshi Population towards Thalassemia Prevention : A Nationwide Study. Res squre. 2020;1–16.

3. Ishfaq K. Assessing Parental Knowledge about Thalassemia. 2017;(January).

4. Ebrahim S, Raza AZ, Hussain M, Khan A, Kumari L, Rasheed R, et al. Knowledge and Beliefs Regarding Thalassemia in an Urban Population. Cureus. 2019;11(7):5–10.

5. Karachi S. Frequency and Awareness of Thalassemia in Families with Cousin Marriages : A Frequency and Awareness of Thalassemia in Families with Cousin Marriages : A Study from Karachi, Pakistan. 2017;(November 2018).

6. Thalassaemia: The present and future for Bangladesh | Daily Star [Internet]. [cited 2022 Jun 1]. Available from: https://www.thedailystar.net/health/news/thalassaemia-the-present-and-future-bangladesh-1738645

7. Hashim S, Inam ALI, Jamil H, Klair N, Nayyar A, Sheikh N, et al. Awareness and Acceptance of Premarital Carrier Screening of Thalassemia among Adults Awareness and Acceptance of Premarital Carrier Screening of Thalassemia among Adults. 2021;(April):2–5.

8. Miri-moghaddam E, Motaharitabar E, Erfannia L, Dashipour A. High School Knowledge and Attitudes towards Thalassemia in Southeastern Iran. 2021;8(1):24–30.

9. Akter K, Khatun S, Hossain MS. Lived Experience of Thalassaemic Children in Bangladesh. Open J Nurs. 2020;10(11):1109–25.

10. Hossain MS, Raheem E, Sultana TA, Ferdous S, Nahar N, Islam S, et al. Thalassemias in South Asia: clinical lessons learnt from Bangladesh. Orphanet J Rare Dis. 2017;12(1).

11. Ishfaq K, Naeem S Bin, Ali J, Ahmad T, Zainab S. Assessing parental knowledge about thalassemia. Pakistan Paediatr J. 2016;40(4):232–6.

12. Uddin M, Ul-Haq F, Sarfaraz A, Khan M, Nazim A, Maqsood B, et al. Frequency and Awareness of Thalassemia in Families with Cousin Marriages: A Study from Karachi, Pakistan. Br J Med Med Res. 2017;21(3):1–11.

13. Kolnagou A, Kontoghiorghes GJ. Advances in the prevention and treatment are changing thalassemia from a fatal to a chronic disease. Experience from a cyprus model and its use as a paradigm for future applications. Hemoglobin. 2009;33(5):287–95.

14. Miri-Moghaddam E, Motaharitabar E, Erfannia L, Dashipour A, Houshvar M. High school knowledge and attitudes towards Thalassemia in southeastern Iran. Int J Hematol Stem Cell Res. 2014;8(1):24–30.

15. Hossain MS, Raheem E, Sultana TA, Ferdous S, Nahar N, Islam S, et al. Thalassemias in South Asia: clinical lessons learnt from Bangladesh. Orphanet J Rare Dis. 2017;12(1).

